# A systematic review of non-clinician trauma-based interventions for school-age children and young people

**DOI:** 10.1101/2023.10.10.23296823

**Authors:** Flo Avery, Natasha Kennedy, Michaela James, Hope Jones, Rebekah Amos, Mark Bellis, Karen Hughes, Sinead Brophy

## Abstract

Exposure to adverse childhood experiences (ACEs) is increasingly recognised as a risk factor for health problems in later life. Awareness of ACEs and associated trauma is increasing in schools and educational settings, as well as demand for supportive services to deal with needs. However, there is a lack of clear evidence for effective interventions which can be delivered by non-clinicians (e.g., the school staff themselves). For this review, we conducted a search across five electronic databases for studies published since January 2013. These studies assessed the efficacy of non-clinician delivered interventions for supporting trauma recovery or improvements in mental health in school age youth (4-18 years) who have experienced ACEs. Out of the 4097 studies identified through the search, 326 were retrieved for full text screening and 25 were included in the final review. There was considerable heterogeneity in study design, outcome measures and the intervention being studied. The majority of studies were assessed to be of weak quality due to convenience sampling of participants and potential bias, indicating there is a lack of high quality research evidence to inform non-clinician delivered trauma-informed interventions for improving mental health outcomes in school-age young people. Cognitive Behavioural Therapy (CBT)-based approaches are tentatively suggested as a suitable target for future rigorous evaluations of interventions addressing ACE-related trauma recovery and mental health improvement in school-age young people.

## Introduction

Adverse Childhood Experiences (ACEs) are potentially traumatic experiences such as child abuse or neglect, or household dysfunction (such as drug and alcohol abuse or domestic violence within the family home), occurring before the age of 18. Young people exposed to ACEs face an elevated risk of unhealthy behaviours and long term physical and mental health problems (1,2). This correlation is particularly pronounced among those who have experienced four or more ACEs (3). While not everyone who has ACEs necessarily experiences a traumatic event (4), the occurrence of multiple ACEs closely predicts the development of trauma symptoms (5).

Promoting ACE awareness and implementing trauma-informed services are increasingly recommended as vital supportive measures, both in healthcare (6) and educational (7) settings. In response, schools and policy-makers are advocating for and embedding trauma-informed approaches (8,9). Schools and educational settings are recognised as crucial settings for providing support to young people affected by potentially traumatic experiences (10).

Numerous reviews have examined the evidence for different systemic approaches and therapeutic interventions for supporting young people with trauma (11–13). Many of the interventions in these reviews are clinical in nature, delivered by psychologists, therapists, or medical and allied health professionals. However, with ACEs being relatively common, for example between 1%-38% (3) reported to experience 4 or more ACE, a more population approach is needed. Yet, there is a notable gap in evidence for interventions suitable for non-clinician delivery (14). This is a barrier for many schools with limited access to clinicians and without mental health professionals on staff. Consequently, teachers and teaching assistants are often tasked with providing mental health support to young people (15,16). While Franklin et al. (17) have suggested that interventions delivered by teachers and non-clinicians can be beneficial for young people’s mental health, they highlight the lack of empirical evidence regarding their effectiveness.

This review, therefore, aims to address the question of what evidence base exists for interventions supporting trauma, specifically those suitable for non-clinician delivery. The objective is to synthesise evidence for interventions appropriate for professionals such as teachers and teaching assistants, acknowledging their increasingly critical role in providing mental health support to young people affected by potentially traumatic experiences.

## Methods

This systematic review was conducted in accordance with the Preferred Reporting Items for Systematic Reviews and Meta-Analysis (PRISMA) guidelines (18). The protocol for the review was registered in the PROSPERO International Prospective Register of Systematic Reviews in June 2023 (ID: CRD42023417286).

### Review Design

We conducted a systematic review of quantitative and qualitative evidence to evaluate the effectiveness of trauma-informed interventions suitable for non-clinician delivery. The study findings were summarised through a narrative synthesis. Heterogeneity in study designs and outcome measures precluded the ability to conduct a meta-analysis. One researcher (FA) extracted study data from full text papers, and this was reviewed by a second researcher (RA). Any inaccuracies were presented to and resolved by a third independent researcher

### Search Strategy

Five electronic databases – Web of Science, Embase, Science Direct, Applied Social Sciences Index (ASSIA) and EBSCO (including CINAHL Plus with Full Text, MEDLINE, APA PsycArticles, APA PsycInfo, Teacher Reference Center, and Education Research Complete) – were searched to identify relevant studies published in English. The literature search was conducted in April 2023. Search terms included: Trauma* OR “Post-Traumatic Stress” OR PTSD and Intervention* OR Treatment* and children OR youth OR young OR adolescen* and education OR school OR teach* OR play. Detailed search terms can be viewed in Appendix 1.

### Eligibility

#### Inclusion Criteria

Articles were included if they met the following criteria: (1) published in English in the last 10 years (e.g since January 2013); (2) reported on any supportive or therapeutic intervention with applicability to trauma recovery, or recovery from the impact of ACEs; (3) intervention was suitable for non-clinician delivery e.g. teacher, teaching assistant or similar; (4) participants were aged 4-18 years (school aged) with any experience of or exposure to ACEs; (5) intervention took place in a school, educational setting, community setting, residential or care settings; and (6) included a validated self-reported mental health outcome relating to trauma or adversity such as post-traumatic stress disorder (PTSD) symptoms as an outcome measure. Both randomised and non-randomised studes were included. Language of publication was restricted owing to constraints of the research team’s capacity. Restrictions were made by publication date to maximise the relevance of studies.

#### Exclusion Criteria

Articles were excluded if they met the following criteria: 1) All participants were over 18 or under 4 years; 2) interventions were assessed indirectly through a teacher or professional assessment of young people; 3) interventions were based on disasters such as earthquakes, floods or experiences of war; 4) interventions were delivered by clinicians; 5) interventions were behavioural rather than supportive, such as violence prevention or sex education programmes; 6) the research study had implemented a case study designs; and 7) the research had been conducted in a hospital or in-patient healthcare setting.

### Study Selection

The search yielded 7147 results, which were imported into Covidence for title and abstract screening. After removing 3050 duplicates, the remaining titles and abstracts (4097) were screened against the selection criteria by the primary author, FA. Additionally, all titles and abstracts underwent independent screening by a second reviewer. Reviewers achieved a 96% agreement rate, and any disagreements were resolved through discussions among authors (FA, SB, NK, RA, HJ, MJ), resulting in a majority consensus reached by agreement between at least three authors. As a result, 3770 studies were excluded as irrelevant. FA retrieved and independently screened all full-text articles (326), which were also independently screened by a second reviewer. Reviewers reached an 88% agreement rate, resolving disagreements through discussion amongst authors (FA, SB, NK, HJ, MJ), with a majority vote from at least three authors. Figure 1 shows details of the selection process, including reasons for excluding articles at different stages. Twenty-four articles met the inclusion criteria. Data extraction from these articles was undertaken by one reviewer (FA) and cross-checked by a second reviewer (RA or MJ). Data extracted included demographic data regarding the papers such as country of study, target population, year of publication, and descriptive information about the intervention such type of intervention, length of followup, measures of effect, barriers to implementation. The outcomes differed in each study and so a descriptive extraction of the data in each study was recorded. The study quality was assessed using validated tools (see below). Furthermore, reference lists from these twenty four studies were manually searched by FA, resulting in the identification of one additional paper. This was uploaded to Covidence and voted on by other authors for inclusion. The paper was voted eligible for inclusion and so a total of twenty-five articles met the inclusion criteria and are included in this review.

**Fig. 1.**
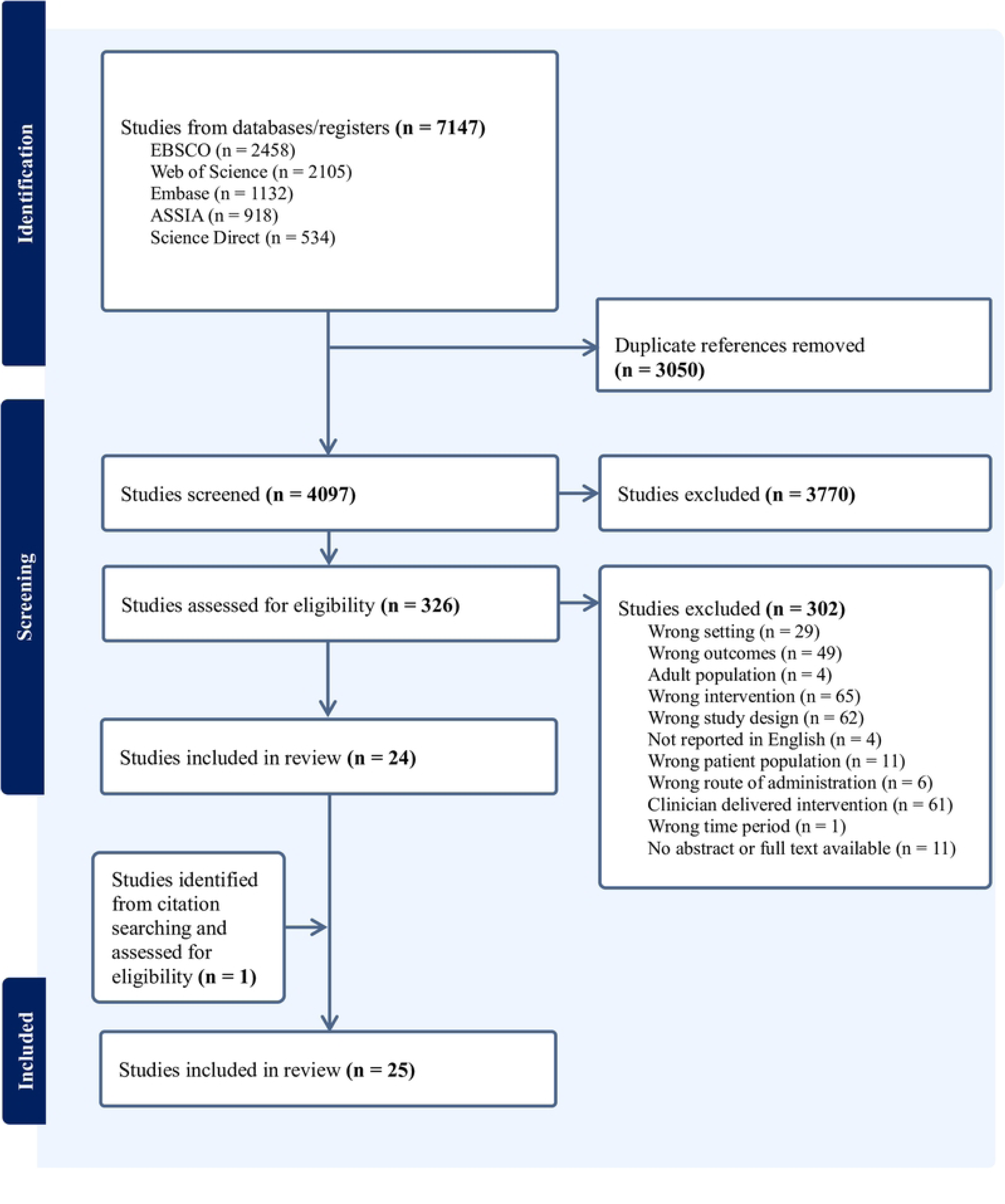
PRISMA diagram.

### Quality Assessment

Effective Public Health Practice Project (EPHPP)’s Quality Assessment Tool for Quantitative Studies was used to assess the quality of quantitative research including randomised control trials (RCTs), non-randomised experimental studies and cohort design studies (19). This tool assesses potential selection bias, the appropriateness of study design, confounding, blinding, data collection tools, participant dropouts, intervention integrity and statistical analyses. Quality ratings are scored on a scale of 1-3, where 1 indicates strong quality, 2 indicates moderate quality, and 3 indicates weak quality. Overall ratings are classified as a) weak (when 2 or more components rated as weak), b) moderate (when only one component rated as weak), or c) strong.

Qualitative studies were assessed using the Critical Appraisal Skills Programme (CASP) Qualitative Studies Checklist (20). This checklist assesses three broad areas, such as a) the validity of the study, b) the study findings and c) the utility of the research locally. Within each of these broad domains are a set of questions designed to provide an overview of the study quality in each domain (20). Mixed methods research was assessed using both checklists. No studies were excluded based on the results of the quality assessment. Quality assessment was conducted by one reviewer (FA) and checked by a second reviewer (RA and MJ). Disagreements were resolved through discussion amongst authors.

## Results

### Overview of studies

Characteristics of the twenty five studies included in the current review are summarised in Table 1. Six studies were randomised control trials (21–26), fifteen used a cohort design (pre-test and post-test) (26–38), three used qualitative methods (39–41) and one used mixed methods (cohort design and qualitative methods) (42). The average sample size was 119 participants, ranging from 15 participants (37) to 565 participants (43). Of the included studies, 14 (56%) had a sample size above 50 (21,27,28,30,32,34–36,39,43–46). As anticipated, there was a high level of methodological diversity in terms of study design, populations (e.g. refugee, LGBTQ, youth offender, middle east, USA, Europe, Asia) and measures used, and therefore meta-analysis was not conducted.

The studies in this review either recruited the study population based on likely exposure to trauma, measured exposure to traumatic events, or measured PTSD symptoms (or a mix of these). Seven studies directly measured exposure to traumatic events (21,22,25,30,35,40,44,47). Thirteen studies directly measured PTSD symptoms (22,24,29,34,40,42), and of these, seven used PTSD score as selection criteria (29,30,33–35,37,43).

Ten studies assessed interventions which were delivered in the school setting (24,25,28–30,33,35,36,38,43). Three studies observed interventions delivered online (22,34,37). Eight studies looked at interventions delivered in a community setting (such as a church or charity site) (21,23,26,27,31,39,44,46), two were interventions delivered on university campuses (40,41), and one was an intervention delivered in a residential setting (32). One study evaluated the intervention across different settings, including a school and residential setting (42).

### Quality of included studies

The quality assessment scores of included studies are shown in Tables 2-4. The methodological quality of included studies was weak overall. Using the EPHPP tool for quantitative studies, two studies were categorised as strong, seven studies as moderate, and the rest as weak. The majority of studies rated as weak relied on convenience samples of participants and did not adequately describe methodological information, such as potential confounding and blinding. Overall, qualitative studies were not considered strong, primarily because they lacked consistent discussions on how the relationship between researcher and participants was managed, including consideration of power dynamics and ethical issues.

The nine studies rated moderate or strong on quality included a range of interventions. Two studies looked at CBT-based groups in refugee populations (29,43). Other studies evaluated interventions focused on expressive writing (32), mindfulness (33), game-based mindfulness (37), trauma-focused CBT (delivered by lay counsellors) (44), a targeted group for sexual and gender minority adolescents (SGMA) (24), and a psychoeducational programme about posttraumatic growth (25).

### CBT-based groups

Seven studies evaluated CBT-based groups, although four were of weak quality. Four studies investigated Teaching Recovery Techniques (TRT), a five week programme based on trauma-focused cognitive behavioural therapy (TF-CBT) designed for children in low resource settings, making it suitable for delivery by non-clinicians. The program was developed by the Children and War Foundation (48). El-Khani et al. (43) evaluated TRT in an RCT with Syrian refugee children aged 9-12 and their families in Lebanon, comparing it with a waitlist control group and a TRT + parenting (TRT+P) condition, which was rated as strong for quality. Data were collected pre-intervention (T1) and post-intervention at 2 weeks (T2) and 12 weeks (T3). All three conditions experienced a significant reduction in PTSD intrusion symptoms measured on the CRIES-13, with the greatest reduction in the TRT+P condition, showing the lowest levels at T3. Both the TRT and TRT+P condition also experienced reductions in depression. Furthermore, all three conditions experienced a significant reduction in anxiety, however the waitlist group’s scores increased by T3. It is suggested that a parenting skills component can enhance the positive effects of TRT. A pilot study by the same authors evaluating TRT in the same population in a Turkish setting was also included (29). This study was rated weak for quality due to selection bias of participants and the absence of a control group. Similar findings were observed; a significant reduction was reported by participants in the Intrusion scores on the CRIES-13 at two weeks post-intervention, although other measures did not reach significance owing to low power (n=16).

Sarkadi et al. (42) evaluated TRT in an adolescent refugee population in Sweden and found significant reductions in PTSD (CRIES-8) and depression (MADRS-S) symptoms two weeks after the intervention. Qualitative data indicated that the programme had utility through provision of social support, normalisation, valuable tools, and manageability. However, there was no reported follow up beyond two weeks; a 3-6 month follow up was planned, but measures were not reported. Barron (26) found similar results in their RCT evaluating the impact of TRT for youth with PTSD in Brazilian favelas, showing a significant reduction in PTSD and depression symptoms, and a small effect size for posttraumatic growth compared with the control group. However, there was no follow up beyond two weeks post-intervention, providing no evidence of long term impact. TRT was the most extensively studied intervention and has the strongest evidence base for reducing PTSD symptoms, although two of these studies were rated as low quality (29,49), and only one followed up beyond two weeks post-intervention (43).

Bryant et al. (21) conducted a moderate-quality RCT evaluating a similar group-based intervention called Early Adolescent Skills for Emotions (EASE) for Syrian adolescents. The intervention spanned seven group sessions focusing on arousal reduction, behavioural activation, and problem management. Data was collected pre-intervention (T1) and post-intervention at 9 weeks (T2) and 3 months (T3). Significant reductions in the PSC-internalising scale were found compared to the control group, and this effect was sustained at T3, although no significant difference were found in other variables. EASE was also evaluated by Akhtar et al. (46) in a pilot RCT, although the study was low powered and did not detect any significant changes.

Elswick et al. (30) evaluated the impact of a group after school club called the Trauma Healing Club, based on Cognitive Behavioral Interventions for Trauma in Schools (CBITS). CBITS is an evidence-based intervention deliverable by a clinician, whilst the Trauma Healing Club is suitable for non-clinicians. While significant improvement in PTSD symptoms was observed, it should be noted that this was a poor-quality study that did not report on withdrawals, the reliability and validity of measures, or potential confounders.

Li et al. (30) assessed the Power up Children’s Psychological Immunity (PCPI) program, a modified group version of TF-CBT. This study was also of poor-quality and did not report on reliability and validity of measures, or potential confounders. Participants reported significantly reduced PTSD, depression, and anxiety following the intervention, but by the 3 month follow up, levels were similar to pre-intervention levels.

Overall CBT-based groups had the strongest evidence base of any intervention type, with three RCTs rated strong or moderate quality providing promising evidence. However it should be noted that the remainder of studies evaluating CBT-based groups were rated weak in quality and are therefore of limited utility.

### Other CBT-based Approaches

Four studies explored CBT-based approaches in three distinct further contexts; one-to-one interventions (44,47), an online platform (34) and an art-based curriculum (38).

Murray (44) conducted an evaluation of TF-CBT for children affected by HIV/AIDS in Zambia. This was a modified lay counsellor-delivered version of the intervention, which is typically administered by clinicians. The sessions involved a combination of individual sessions with the child, caregiver(s) separately, and family sessions. Lay counsellors, lacking formal qualifications in counselling or mental health, typically provide psychological support through a community setting, often with a faith-based focus. This study received a moderate quality rating, revealing significant reductions in the severity of trauma symptoms and shame symptoms when comparing pre-intervention and two weeks post-interventionscores. However, there was no long term follow up. The authors did not provide a detailed description of how TF-CBT was adapted for non-clinician delivery, but it included cultural adaptations and simplified terminology. Close weekly monitoring was provided by the trainer and a supervisor to ensure fidelity.

Durmoney et al. (47) evaluated the impact of a peer delivered trauma-informed CBT skills curriculum among young men aged 17-24 years involved in the justice system. The intervention was delivered by ‘Paraprofessionals’ such as youth workers and volunteers in community settings, often within participants’ homes or on the street, to maximise engagement. This study was of weak quality, with a high likelihood of selection bias and unreliable or unverified data collection tools. Although there were indications of significant improvements in distress related to employment and education over time, emotion regulation did not show significant improvement.

Jaycox et al. (34) assessed an online stress and trauma curriculum called Life Improvement for Teens (LIFT), incorporating elements resembling TF-CBT and CBITS. The self-paced, internet-based curriculum was delivered online to high school students, who accessed the intervention during a weekly slot at the school site. LIFT comprised both a stress track and a trauma track, with students who reported potentially traumatic experiences being allocated to the trauma track. This study found significant improvements in PTSD symptoms and emotional problems, but observed no change in depression or anxiety. School functioning deteriorated over the course of the study. Overall, this study was rated as weak quality, as it did not specify the validity of data collection tools, and it did not discuss reasons for participant withdrawal.

Sitzer and Stockwell (36) also evaluated an art programme which incorporated aspects of CBT, which is discussed in the subsequent Art and community-based approaches section.

### Mindfulness

Three studies focussed on evaluating a mindfulness intervention (22,33,37). Ito et al. (33) assessed the effects of short-term mindfulness-based group intervention delivered within a school setting for adolescents with trauma. Participants attended a single 60-minute group session and were subsequently encouraged by teachers to continue using mindfulness skills and cognitive defusion techniques. The study revealed significant improvements in mindful attention and awareness, along with reductions in depression, anxiety symptoms, and posttraumatic stress symptoms related to hyperarousal. However, these measures were recorded two weeks after the intervention, thus long term effects are unclear. This study was rated moderate quality. Schuurmans et al. (35) compared the impact of three meditation-based online games among youth in a residential care setting. Participants engaged in twelve 15-minute sessions twice a week for 6 weeks. One game, called Muse, demonstrated improvements in post-traumatic symptoms, stress, and anxiety at the one month follow-up, while results for the other games were inconsistent. Although the study had a moderate overall quality rating, it featured a very small sample size, with n=15 at randomisation and n=9 at the one month follow up. Davis and Aylward (22) evaluated the impact of a trauma-informed mindful yoga intervention, which was delivered remotely to high school students. However, the study did not find significant differences in depression, anxiety, or resilience following the intervention.

### Art and community-based approaches

Five studies evaluated interventions based on art, culture, or community, including two cohort design studies (27,38) and three qualitative studies (39–41). All studies received a weak quality rating. Sitzer and Stockwell (38 assessed a Wellness Programme delivered in schools, drawing on elements of art therapy along with aspects of CBT and Dialectical Behavioural Therapy (DBT). Results indicated significant increases in resilience and social and emotional functioning, particularly among male students. However, only unvalidated measures were used in this study. Barnett et al. (27) assessed the impact of residential culture camps on the wellbeing of Alaska Native youth aged 13 to 18 years. The results suggested a significant increase in positive affect, self esteem, and a sense of belonging. However, not all measures in this study were validated.

Harden et al. (40) examined the effects of a twice weekly after school programme involving media and theatre activities as youth-led empowerment strategies, conducted on a university campus. The study identified positive themes related to empowerment, post-traumatic growth, and peace restoration among participants. Nevertheless, there were numerous methodological issues. The stated aim of the study was ‘to contribute to a narrative that would reflect the TNT project’s value’, which is biased. There was no consideration of the relationship between researchers and participants, or ethical issues. These methodological weaknesses were common across all included qualitative studies.

Özden Bademci et al. (41) also evaluated a programme held on a university campus, featuring creative workshops and mentoring from volunteer university students for homeless boys aged 14-17 years. Qualitative themes indicated that participants perceived the campus as a safe, engaging environment, developed trusting relationships with mentors, and increased their capacity for emotional regulation. However, similar methodological critiques applied, including a lack of consideration for the potential ethical issues arising from the mentoring relationships, and insufficient detail provided concerning the interview protocol. McMahon & Pederson (39) assessed a community-based juvenile justice diversion program and identified positive themes related to self-efficacy, empathy, connection, and conflict resolution. However, the research protocol, including how themes were reached, was not described. Overall, the evidence for art and community-based approaches found to be very weak.

### Other interventions

The remaining interventions, which did not fit into thematic categories, will be summarised individually. Greenbaum & Javdani (32)2 evaluated a therapeutic writing intervention called Writing and Reflecting on Identity To Empower Ourselves as Narrators (WRITE ON), delivered to juvenile justice-involved youth. The intervention consisted of 90 minute sessions twice a week for six weeks. The study observed significant increases in positive mental health attributes and resilience throughout the programme and at a two week follow up. The quality was strong, although the long term impact remained unclear.

Goldbach et al. (24) conducted an RCT to assess a 10 week intervention called Proud & Empowered (P&E), which targeted SGMA and was conducted during the school day. Participation in the P&E intervention reduced minority stress and improved depression and suicidality measures, although there was no significant change in PTSD symptoms. Data was collected only one week after the intervention finished. This study received a moderate quality rating overall; validity and reliability of measures was not stated.

Taku et al. (25) examined a pychoeducational intervention about posttraumatic growth (PTG) in three conditions: a Control Group, Group 1 (intervention focused on stress-related reactions and PTG), and Group 2 (focused on negative changes and PTSD only). In Study 1, Group 1 and the Control Group showed higher PTG than Group 2 after two weeks, suggesting that exposure to negative information about PTSD may supress PTG. In Study 2, there were no differences in PTG perceptions between the groups. Findings were inconclusive for this moderate quality study.

Eruyar and Vostanis (31) evaluated the impact of group Theraplay, an attachment based intervention, with refugee children and their parents. After eight weekly sessions, a significant improvement was found in PTSD scores. However, only 59% of participants completed the programme, and reasons for withdrawals were not provided. Additionally, not all data collection tools were validated, although the PTSD measure was, which limited the quality. This study was one of five studies, of varying quality, assessing interventions involving both children and caregivers, all of which suggested a positive impact (29,31,42–44).

Day et al. (28) assessed a trauma-informed intervention in a residential school, involving curriculum changes and access to relational interventions such as Theraplay to build attachment, self-esteem, and trust in others. Provision was made for two rooms to act as alternative environments for struggling students, providing access to problem solving, talk therapy, and use of sensorimotor activities. Significant differences were found in the pre and postest scores for posttraumatic symptoms. However, the study was weak in quality given the high chance of selection bias and a lack of validated measures. Furthermore, the intervention was broadly described without specific details.

Martin and Wood (36) evaluated a group drumming intervention delivered in a school and found significant improvements for boys in terms of higher mental wellbeing and lower post-traumatic stress symptoms, but not for girls. This study was of weak quality due to selection bias and a high unexplained drop out rate.

## Discussion

A systematic review methodology was used to investigate the evidence base for interventions designed to support young people affected by trauma and are suitable for delivery by non-clinicians. This is the first systematic review to comphrensively appraise interventions in this domain specifically designed for non-clinician delivery. Overall, the evidence base for trauma recovery interventions was found to be weak. Only nine out of the twenty-five studies identified were rated as strong or moderate quality, and among these, only seven reported significant improvements in mental health outcomes, including depression, anxiety, or PTSD (21,26,32,33,37,43,44). Among these seven, four adopted a CBT-based approach (21,26,43,44), while two followed a mindfulness approach (33,37). The most frequently studied CBT-based intervention was TRT (26,43,49). A number of studies evaluating TRT were excluded earlier in the review process because they were administered by a clinician. While TRT itself is suitable for non-clinician delivery, some excluded studies involved clinician delivery to enhance fidelity. CBT-based groups, particularly TRT, are tentatively suggested as the interventions in this domain with the strongest evidence base.

Several of the included studies trialled interventions with refugee populations (21,29,30,43,46,49,50). This area has a growing evidence base, considering that refugees are often supported in low-resource environments such as refugee camps, where accessing specialised support can be challenging even after achieving settled status. Notably, this review excluded studies specifically measuring war- or disaster-related trauma. As a result, the studies which have been included from this context are more generalisable to a wider range of young people affected by adversity.

Six studies evaluated interventions which involved caregivers in some or all of the sessions which formed the intervention (29,43,44,46,49,50). El-Khani et al. (43) observed that TRT achieved a greater positive impact on young people’s mental health and PTSD symptomology when a parenting component was included. This finding suggests a potential avenue for future research, given the intergenerational nature of ACEs and the recognised role of trusted relationships with close adults as a potential protective factor for young people exposed to ACEs (51).

### Implications for research

In this domain, there is a notable scarcity of high quality studies, highlighting the need for further RCTs to assess the potential impact of interventions. CBT-based groups and mindfulness interventions appear to be promising targets for future research. Rather than relying solely on clinician-delivered designs to ensure fidelity of interventions intended for non-clinicians, more robust results can be derived from studies that implement non-clinician delivery, supplemented with measures such as observation or clinical supervision to maintain fidelity. Additionally, it is essential to incorporate longer-term follow up into study design. A majority of studies included in this review collected post-intervention data within a period of 2 weeks or less. To establish the robustness of intervention evidence, longer-term follow up assessments are imperative.

In situations where conducting RCTs may not be feasible, efforts should be directed towards ensuring the reliability and validity of data collection tools. Furthermore, researchers should diligently record and follow up on withdrawals from the study. This practice is both methodologically and ethically important, as traumatised youth may disengage from therapeutic interventions if they perceive them as harmful or unsupportive. Exploring this perspective can provide valuable insights into improving intervention strategies.

### Implications for educational settings

There is not a strong, definitive recommendation for schools seeking to adopt a research-informed approach within their educational setting to support young people who have experienced trauma and adversity. However, based on available evidence, the approaches which are most likely to be effective at supporting young people with trauma recovery are CBT-based groups or mindfulness approaches. Additionally, interventions involving caregivers may also have value in this context. This aspect might be particularly pertinent for primary schools, which serve pupils aged under 11 years in the United Kingdom, and often maintain closer partnerships with parents and caregivers.

### Review limitations

The overall quality of this systematic review was assessed using AMSTAR 2 (52). The review meets or partially meets all checklist items on AMSTAR 2. Although the time frame for follow up was not stated in the inclusion crtieria, it has been reported for all strong and moderate quality studies. The review did not examine study registries and did not seek to consult experts in the field and sources of funding for the studies included in the review have not been reported. Given the high level of anticipated and actual heterogeneity of studies in this area, a meta-analysis was not planned or conducted, which restricts the scope of the conclusions that can be drawn. Finally, the work only examined studies from the past ten years, only selected those published in English, and may have missed those published in non-academic literature. The secondary screen of the references of papers may have help to identify some of the articles missing due to the search criteria, but future work would be needed to idenifty interventnions published in grey literature or published in lanuages other than English.

## Conclusions

Our review of 25 studies reveals inconsistent evidence regarding non-clinician delivered trauma informed interventions for enhancing mental health outcomes in school-age individuals. There are a number of strategies with evidence that would suggest more development and rigorous evaluation would be of benefit. That is especially including an economics or cost effectiveness evaluation of those delivered online or further evaluation of the CBT-based approaches. As schools and educational institutions are increasingly expected to assume a greater role in supporting mental health of traumatised youth, the development of this evidence base, is of crucial importance.

## Avaliablity of materials

Data collection templates, the Covidence search results and any other materials are available upon request to.

## Author Contributions

Flo Avery led the review, was responsible for managing the project, analysing data and preparing an original draft.

Michaela James contributed to abstract and full text screening, quality assessment and editing.

Rebekah Amos contributed to abstract and full text screening, and quality assessment, editing and writing.

Natasha Kennedy contributed to abstract and full text screening, and editing.

Hope Jones contributed to abstract and full text screening, and editing.

Sinead Brophy, Karen Hughes and Mark Bellis provided supervision and advice for the systematic review and methodology, and reviewed the final report.

## Data Availability

This work is based on publicly available papers and so is in the public domain.

